# A roadmap to account for reporting delays for public health situational awareness – a case study with COVID-19 and dengue in United States jurisdictions

**DOI:** 10.1101/2024.11.09.24315999

**Authors:** Velma K. Lopez, Leonardo S. Bastos, Cláudia T. Codeço, Michael A. Johansson

## Abstract

**Background:** Decision-making in public health is limited by data availability where the most recent reports do not reflect the actual trajectory of an epidemic. Nowcasting is a modeling tool that can estimate eventual case counts by accounting for reporting delays. While these tools have generated reliable predictions when designed for specific use cases, several limitations exist when scaling the models to systems composed of multiple distinct surveillance systems. We seek to identify flexible application of nowcasting models to address these problems.

**Methods:** We used a previously developed Bayesian nowcasting tool, which dynamically estimates delay probabilities up to a user-defined maximum delay using a user-defined training window. We tested automated approaches to select the maximum delay and training window, setting maximum delay values at the 90^th^, 95^th^, and 99^th^ quantile distribution of the most recently reported data and training windows to the maximum delay plus one week or multiplied by 1.5 or 2.0. We generated and evaluated 321 nowcasts for COVID-19 cases in six U.S. states and dengue cases in Puerto Rico. We assessed prediction error and precision via logarithmic scoring and coverage metrics for the most recent three weeks of predictions in each nowcast. We used these metrics to further assess why nowcasts may fail and to compare predictions generated from three different publicly available tools.

**Results:** Using recent data to estimate dynamic delay and training window parameters resulted in nowcast with less error relative to nowcasts made with static parameters for long historic periods. Nowcasts likely to fail could be predicted *a priori* by the relative width of the prediction intervals and the permutation entropy of the epidemic trend. More complex models do not necessarily improve performance, for example, a model with random effects for reporting periodicity did not improve nowcasts compared to a simple model which fit the observed epidemic trend.

**Conclusions:** We tested multiple systems for scaling up nowcasts in a flexible framework. We recommend using dynamic parameter selection and creating a system to suppress nowcasts likely to fail. This requires collaboration with surveillance colleagues to implement data-driven choices to improve the utility of predictions for decision-making.

## Introduction

Reporting delays are inherent in all public health surveillance systems. The time between symptom onset and when an illness is captured in a public health agency database can be influenced by many factors, including, but not limited to, disease presentation, care seeking, testing protocols, and reporting processes (1). Consequently, when tracking the number of reported cases by onset date over time, the epidemic trend in the most recent weeks will always appear to decline, regardless of the true underlying trend. Without some sort of adjustment to the trend, raw surveillance data may have limited interpretability for timely public health action (2). Nowcasting, that is, a quantitative method to predict the number of cases that have occurred but are not yet reported, is one approach for reducing the impact of reporting delays on situational awareness.

While not yet a routine component, nowcasting has been increasingly integrated into public health surveillance and outbreak response. For example, since 2015, the Brazilian government has incorporated nowcasting into public municipality-level communication tools for dengue, chikungunya, and Zika (https://info.dengue.mat.br/; (3)). These dashboards are designed to inform public health action based on quantified levels of current transmission risk. In the past few years, nowcasting has also been integrated into analyses for the recent United States (US) federal government and local response to mpox (4,5), and at the state (6) and local level (7) during the COVID-19 pandemic. Moreover, researchers throughout the world have worked with government partners to integrate nowcasting in COVID-19 response in Bavaria (8), Germany and Poland (9), and Sweden (10), and mpox in the United Kingdom (11). Their implementation differed, each using different approaches to fitting time series – some groups included random time effects while others augmented models with additional data sources. Importantly, local health department authors have called on the scientific community to create practical nowcasting implementation guidance (5), including model optimization and evaluation. Central to this discussion is how to select model parameters within the simplest modeling structure that yields the most accurate predictions.

Despite the increasing application of nowcasting in public health, the existence of several out-of-the-box nowcasting tools, and the call from health departments for practical guidance, many barriers prevent jurisdictions from implementing nowcasts as part of routine public health. This is in part due to the many challenges inherent in nowcast model building. First and foremost, small fluctuations in reported cases or changes in reporting delays can manifest large prediction uncertainty. Second, because precision is challenging, detecting change points in transmission (e.g., a peak or a sudden increase in cases) is also difficult. Many jurisdictions thus suppress the most recent weeks of cases to avoid these modeling problems. These latest data, however, can provide meaningful insights since a nowcast can capture important uncertainty that may not be immediately apparent. Nonetheless, optimizing certainty to be properly calibrated is critical. Stable nowcasts depend on a tradeoff between acquiring robust estimates of delays with longer training periods and delay windows versus being able to dynamically respond to changes in reporting by using short training periods. For example, too long of a training period may result in a smoothed trend, and a change in reporting may be missed. However, updating model parameters manually is not feasible at scale, for example, for many locations with weekly nowcasts, necessitating either fixed or algorithmic approaches to select key parameters. Moreover, it is also imperative for practitioners to select appropriate methods and assess when the predictions may be misleading and when their use for decision-making may cause more harm than good. This requires validation and comparison of methods both prospectively and retrospectively, to ensure that nowcasts are performing as expected and to encourage cautious use at times when they may be misleading, such as during non-routine reporting delays. Here, we seek to assess nowcast performance for varying epidemiologic surveillance systems to guide implementation across diverse systems. Our goals are to 1) identify an optimized process for implementing existing nowcast tools when historical data are limited and reporting delays vary; 2) better understand and identify scenarios in which nowcasts fail; and 3) identify trade-offs in different nowcasting models.

## Methods

### Surveillance systems data

To assess nowcast performance in varying surveillance systems, we fitted models to both epidemic and endemic diseases. Surveillance system delays for epidemic diseases generally exhibit heterogeneity in reporting patterns as well as disease magnitude. Analysis of such systems may provide insight into nowcasting performance under conditions of evolving epidemiology. In contrast, important insight may also be gained by analyzing reporting patterns for endemic diseases. Surveillance systems for endemic diseases are generally well-established and exhibit timely reporting. These systems may also include expected periods of lower burden or sporadic occurrence of cases.

For epidemic surveillance, we used COVID-19 cases reported from U.S. jurisdictions to the Centers for Disease Control and Prevention (CDC). We selected a subset of state-level jurisdictions based on data completeness and variability of reporting. We first excluded all jurisdictions that did not report cases for at least 90% of weeks in the analysis period (n=13). The remaining jurisdictions were categorized based on the mean and standard deviation of reporting delays greater than 91 weeks (July 5, 2020 – March 26, 2022; see Supplemental Figure 1 for distribution of reporting delays in all jurisdictions) into four groups: short or long delays (mean reporting delay greater than or equal to 4 weeks) and low or high variation in delays (standard deviation greater than or equal to median standard deviation across all jurisdictions, see Supplement 1). We excluded all jurisdictions classified as having a long mean delay and low variation in delays because many of these jurisdictions had several months with extreme delays that were more likely artifacts than true reporting delays (for example, Missouri, Minnesota, and Texas) while others had sparse data (such as West Virginia). We consider these states to be outliers and hypothesize that their inclusion in the analysis may lead to spurious findings. In order to limit computation time, two jurisdictions were randomly selected per group and the following jurisdictions were included in the main analysis:

- Florida and Michigan from the long mean delays with high variation group,
- North Carolina and Wisconsin from the short mean delays with high variation group,
- Idaho and South Carolina from the short mean delays with low variation group.

To assess performance for an endemic disease with an established surveillance system, we also analyzed dengue virus cases in Puerto Rico (213-week period, selecting every 4^th^ week from January 1, 2010 - December 31, 2013). Dengue cases are reported to the Puerto Rico Department of Health.

All data were aggregated to the weekly scale. Cases with erroneous negative reporting delays were excluded from further analysis. For each of the seven datasets analyzed, we sequentially ran nowcasts moving forward in time with the first nowcast for week 12 in each data set, including all data reported by that week in that dataset. We then stepped forward one week at a time for COVID-19 and four weeks at a time for dengue, stopping at 8 months before the last reported case date in order to allow for retrospective performance evaluation. In total, this resulted in 321 nowcasts models on sequentially expanding datasets (n=280 for COVID-19 in the 6 states and n=41 for dengue in Puerto Rico) with unique ending dates, which we refer to as the “target date” for each nowcast.

### Nowcasts

We fitted negative-binomial nowcast models using the NobBS R package. NobBS is a Bayesian model that accounts for reporting delays estimated from recent data (12). It was originally designed to nowcast influenza data. Briefly, NobBS fits a log-linear model to reported cases at each time step with random effects for time and the probability of the reporting delay. To reflect the autoregressive nature of an epidemic, reported cases have a first-order random walk. Default priors on the reporting delays are weakly informative to reflect an equal probability of reporting across the range of observed values. NobBS requires two parameters: (i) maximum delay, the maximum length of delays to be estimated, and (ii) training period, the period over which to estimate delays (called ‘window period’ in the function). By default, the function sets the maximum delay to the total number of weeks in the dataset minus 1 and the training period to the total number of weeks in the dataset. We selected fixed and dynamic maximum delay and training period parameters (see below). We ran the model with default package settings – one chain per model at 10,000 iterations each, with a burn-in period of 1,000 iterations – and used default settings for model priors. The package’s standard output does not include convergence metrics, so we suppressed nowcasts that were indicative of failure from the evaluation (see “Failed nowcasts” below).

### Fixed and dynamic parameters

We fitted NobBS models under two different fixed parameter scenarios: (1) a fixed maximum delay of 12 weeks and training period of 24 weeks, and (2) a fixed maximum delay of 24 weeks and training period of 48 weeks. We also tested empirical methods for selecting these two values from the data selected dynamically in two steps:

1. We selected an initial maximum delay value to correspond to the 90^th^, 95^th^, and 99^th^ quantile of the distribution of onset-to-report delays for the most recent week of reported data, enforcing a minimum value of 4 weeks and a maximum value 12 weeks. Quantiles were selected based on empirical observation.
2. We used a series of multipliers to adjust the maximum delay and training period values relative to the value from Step 1. For the maximum delay, we tested multipliers of 1.0 (i.e., no change) and 2.0. For the training period, we used values corresponding to the maximum delay plus one week or multiplied the maximum delay value by 1.5 or 2.0. We also included maximum delay multipliers of 3.0 with a window multiplier of 3.0.

When selected values were longer than the available history, the entire history was used (i.e., the default approach).

### Evaluation

Predictive performance was assessed via logarithmic (log) scoring and prediction interval coverage on the last 3 predicted counts (i.e., the last three weeks in the dataset). We focused on the last three predicted counts because this period is when delays are common and most substantial. For each nowcast, we computed the mean and dispersion of the predicted nowcast samples. Using these values, we then calculated the probability that the eventually observed outcome fell within the distribution of nowcast samples. The logarithm of this probability is the log score, with high log scores (closer to zero) indicating a greater probability assigned to the observed outcome and better performance. We used a lower bound probability of 1 per 10,000 to classify nowcasts as “failed” (see below). We calculated the proportion of “failed” nowcasts for each model according to this criterion. We then removed all nowcasts where at least one nowcast model “failed” (44% of all created nowcasts) and calculated log scores on a consistent set of observed values for all models.

Prediction interval coverage was calculated by determining the frequency with which the 50% or 95% prediction interval (i.e., the credible interval) contained the eventually observed outcome. Jurisdictional-level performance metrics are presented in Supplement 2.

### Failed nowcasts

As described above, we considered a nowcast to have “failed” when there was a mean probability of less than 1 per 10,000 of observing the number of reported cases that were eventually reported (log score = -9.2) over the last three weeks of the nowcast. Log scores are correlated with case counts (13) and we expect some nowcasts to “fail” due to high case numbers and not only poor nowcasts.

We assess components of “failure” that could be used to suppress a nowcast through a robust, yet hypothesis-generating, data reduction approach. We used a random forest classifier, a method shown to have improved generalizability relative to other techniques, and is appropriate for correlated data. Specifically, we trained a 1,000-tree random forest classifier with the following features (see Supplement 3 for correlation between them):

- The mean number of reported cases in the last three weeks;
- Width of the nowcast 50% prediction intervals and the 95% prediction intervals (i.e., the lower limit subtracted from the upper limit), each relative to the mean cases reported in the last three weeks; and
- Stability of the epidemic trajectory, measured via permutation entropy of the entire history prior to the nowcast date; and permutation entropy of the 12 weeks prior to the nowcast date. Permutation entropy values range between 0 and 1, where larger values are indicative of epidemic trends with higher complexity that we refer to as unstable because there is more irregularity in the trend. Conversely, we consider epidemic trajectories with lower permutation entropy to be stable.

We chose these features to both adjust for drivers of high log scores (the magnitude of cases) and because they are available at the time the nowcast is created.

We used the default number of random features as candidates (square root of total number of features) for each split during tree generation and selected the most important determinants of nowcast failure from the random forests’ permutation-based variable importance scores. The scores represent the mean decrease in classification accuracy over the entire forest when the features are removed from each tree. The change in accuracy is estimated by using the sub-sample of data that was not used to build a given tree. We trained the algorithm using 80 percent of the dataset; the remaining 20 percent was used to evaluate model accuracy, sensitivity, and specificity. We also assessed the predictive capability of using the indicators selected from the random forest classifier with a logistic regression model. Florida nowcasts were excluded from this sub-analysis given the extreme log-scores of predictions (99% of nowcasts were classified as “failed”).

### Model comparison

After evaluating parameter selection approaches with NobBS, we used the optimized parameter set to fit negative-binomial nowcasting models with nowcaster (14) and epinowcast (15) using COVID-19 data for two representative locations: Idaho and Michigan. We compared model performance across all three models to assess how differences in the models influence performance. Like NobBS, nowcaster also uses a Bayesian log-linear model to predict reported counts by fitting a multilevel model with reported cases at each time step. Similarly, epinowcast also has an autoregressive model of counts but provides slightly more flexibility by allowing users to change the distribution of reporting delays. Epinowcast also extends the basic framework underlying nowcaster and NobBS to include a model for reporting patterns by weekday or week. In theory, users may benefit from implementing epinowcast if they feel that there are distinct delays related to the onset and reporting. The packages use distinct computational approaches for the nowcast function: NobBS and epinowcast use Markov chain Monte Carlo (MCMC), whereas nowcaster uses the Integrated Nested Laplace Approximation (INLA). Additional details on model differences are presented in Supplemental Table 4.

For nowcaster models, we used the default value of 1,000 samples drawn from the approximate posterior distribution. For epinowcast models, we ran eight chains with 1,000 iterations and a burn-in period of 250 samples. Supplemental Table 5 provides a summary table of the similarities and differences between these models. At the time of this analysis, epinowcast could only be used to estimate daily predictions; we aggregated daily samples to the weekly scale for comparison with predictions from other models. Gelman-Rubin statistic (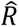), a convergence diagnostic, and model run-time are presented in Supplement 4.

Each model is configured to output prediction intervals rather than parametric forecasts. We therefore assessed performance based on prediction interval coverage and relative weighted interval scores (rWIS) on the last 3 predicted counts. WIS reflects a weighted estimate of sharpness (i.e., the range of the predicted interval) and calibration (i.e., magnitude of over or under prediction) across three prediction intervals (50%, 95%, 98%) and the median prediction. To calculate rWIS, we first estimated the geometric mean of WIS across predictions and models in order to estimate a standardized rank score, and then divided that value by the WIS of the NobBS model. Relative WIS values greater than 1.0 had worse performance than NobBS, and values below 1.0 reflected better performance.

Analyses were conducted using R (version 4.4.0) (16) on a 64-bit Windows 10 desktop (128 GB RAM, Intel(R) Xeon(R) w5-3433 with 1.99 GHz processor). The following packages were used for primary analyses: *NobBS* (17), *nowcaster* (18), *epinowcast* (15), *statcomp* (19), *scoringutils* (20), and *randomForest* (21). R code is available in a public repository (https://github.com/cdcepi/Nowcasting_scale_up_analysis).

This activity was reviewed by CDC to be deemed not human subject research and was conducted consistent with applicable federal law and CDC policy^1§^.

## Results

### Reporting delays

We selected seven unique surveillance systems as a test bed for assessing nowcast performance. Reporting delays across these systems had a median of 2 weeks, interquartile range of 5 weeks, a mean of 5.6 weeks, and a standard deviation of 8.7 weeks, with substantial heterogeneity between systems (Figure 1). For example, in the datasets used here nearly all COVID-19 cases in Florida in the fall and winter of 2020 had reporting delays of at least 15 weeks and 10-25% of COVID-19 cases occurring in Michigan in the spring of 2021 were reported nearly a year after they occurred (Figure 1A). Some of these represent extended delays for backfill and others represent retrospective batch reports. For example, in the Florida nowcast in Figure 1C, there was a median case count of 14 cases over the last three weeks at the time of the nowcast, but a median of 22,954 cases was eventually reported for those dates. More than a quarter of this analysis (26% of included weeks) had a mean difference in reported cases greater than 20,000 cases, suggesting large batches of cases were reported several weeks after they occurred. Excluding those time periods and others with particularly long delays in Michigan and Wisconsin, we found that most delays were less than 4 weeks, the minimum value we allowed for the maximum delay parameter (Figure 1B).

**Figure 1:**
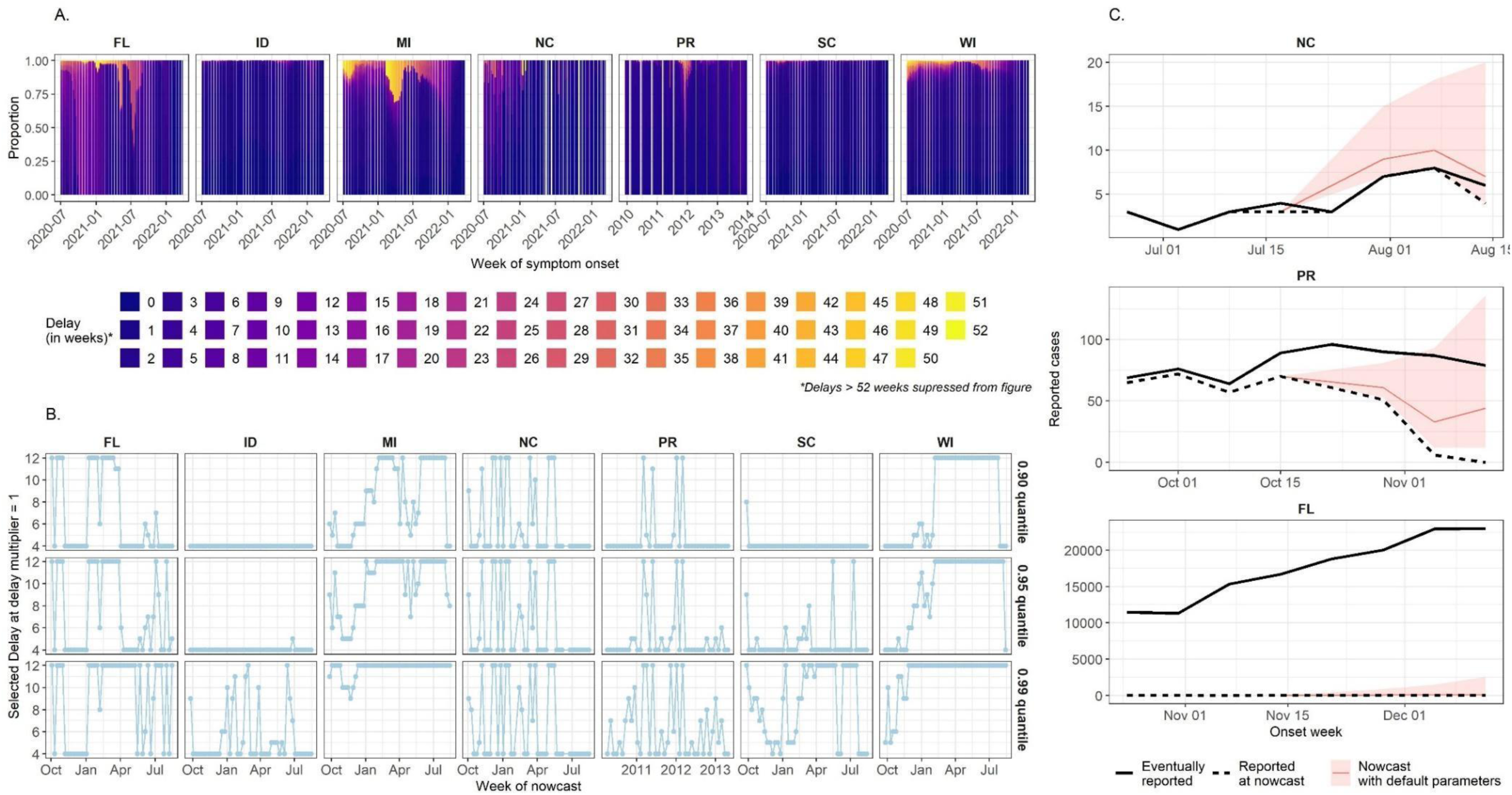
A. Distribution of delays across all surveillance systems by onset week, with each vertical bar representing the delay distribution within that given onset week; B. Dynamically selected delay values for delay multiplier 1 for each quantile value across all surveillance systems; C. Examples of three nowcast predictions, with the median prediction in red and 95% prediction interval in the red band. Reported cases are also presented, with cases reported at the time the nowcast was made in the black dashed line and cases that were eventually reported in the solid black line.

### Nowcast performance

In the selected jurisdictions, we ran nowcasts with default, fixed, and dynamic parameters and assessed predictive performance of each parameter set with log scores and prediction interval coverage on the last 3 predicted counts. Once nowcasts were scored, we classified nowcasts as “failed” if they had scores of less than -9.2 (i.e., if the nowcast probability of the observed outcome was 1 in 10,000 or less). Across parameter sets, 19% to 31% of nowcasts failed by this criterion, and failure was more common with the longer maximum delay values (Figure 2A). The log scores of non-failed nowcasts had similar interquartile ranges across all parameter sets, though the default parameters performed worst, with the lowest mean (-6.3) and median (-7.1) log scores (Figure 2B). Despite performing worst overall, the default model outperformed the dynamic parameter models approximately 20% of the time in PR and up to 10% of the time in NC (Supplemental Figure 2.1).

**Figure 2:**
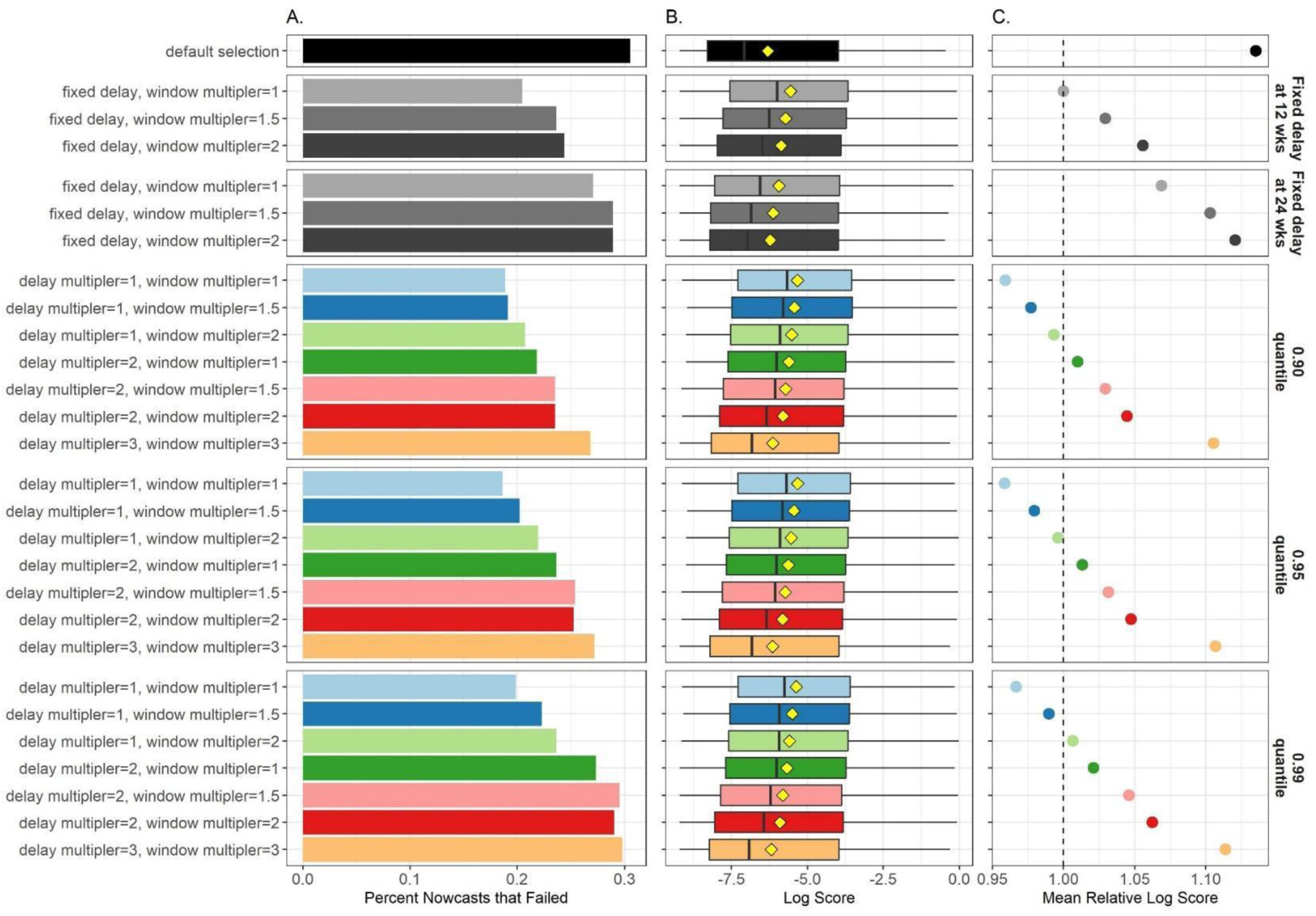
A. The percent of nowcasts with failed log scores, which were excluded from the evaluation; B. Median log score (as a vertical line), interquartile range log scores (as a horizontal bar), and mean log score (as a yellow diamond); C. The parameter set mean log score relative to mean log score of the fixed delay of 12 weeks with 1 window multiplier. In each plot, the parameter set is presented in a distinct color. Nowcasts for dates that “failed” for any model were excluded from all models in panels B and C.

Longer maximum delay parameters were also associated with lower log scores. The fixed 12-week maximum delay outperformed the default parameters (Figure 2C). Extending the 12-week delay or the training window led to worse performance. The dynamically selected parameters were shorter and generally outperformed the 12-week fixed maximum delays, especially without multipliers for the maximum delay or training window. Within the different quantiles used for setting the dynamic delays, extending the maximum delay or training window led to worse performance. Overall, the lowest proportion of failed nowcasts and the highest mean log score was attained with the maximum delay set dynamically to 95^th^ quantile of the onset-to-report delays and the training period set to the maximum delay plus 1. The patterns across surveillance systems largely also reflect the patterns for each surveillance system, with the 95^th^ quantile dynamics delay with no multiplier having the highest or second highest log score despite a slightly higher than average “failure” rate for dengue in Puerto Rico (Supplemental Figure 2.2).

We also assessed prediction interval coverage to evaluate the reliability of the uncertainty estimates from the nowcast. All the models had lower than nominal coverage values for the 50% or 95% prediction intervals (Figure 3). Coverage followed the general patterns of the log scores, with the default parameter selection having the lowest coverage values (37% and 84%, respectively). However, coverage was not maximized for the model with the best log score; doubling the maximum delay parameter for the 95^th^ quantile delay led to higher coverage (45% vs. 49% and 84% vs. 90%, respectively). Coverage varied substantially by jurisdiction, but the trends in relative coverage across models were consistent (Supplemental Figure 2.3).

**Figure 3:**
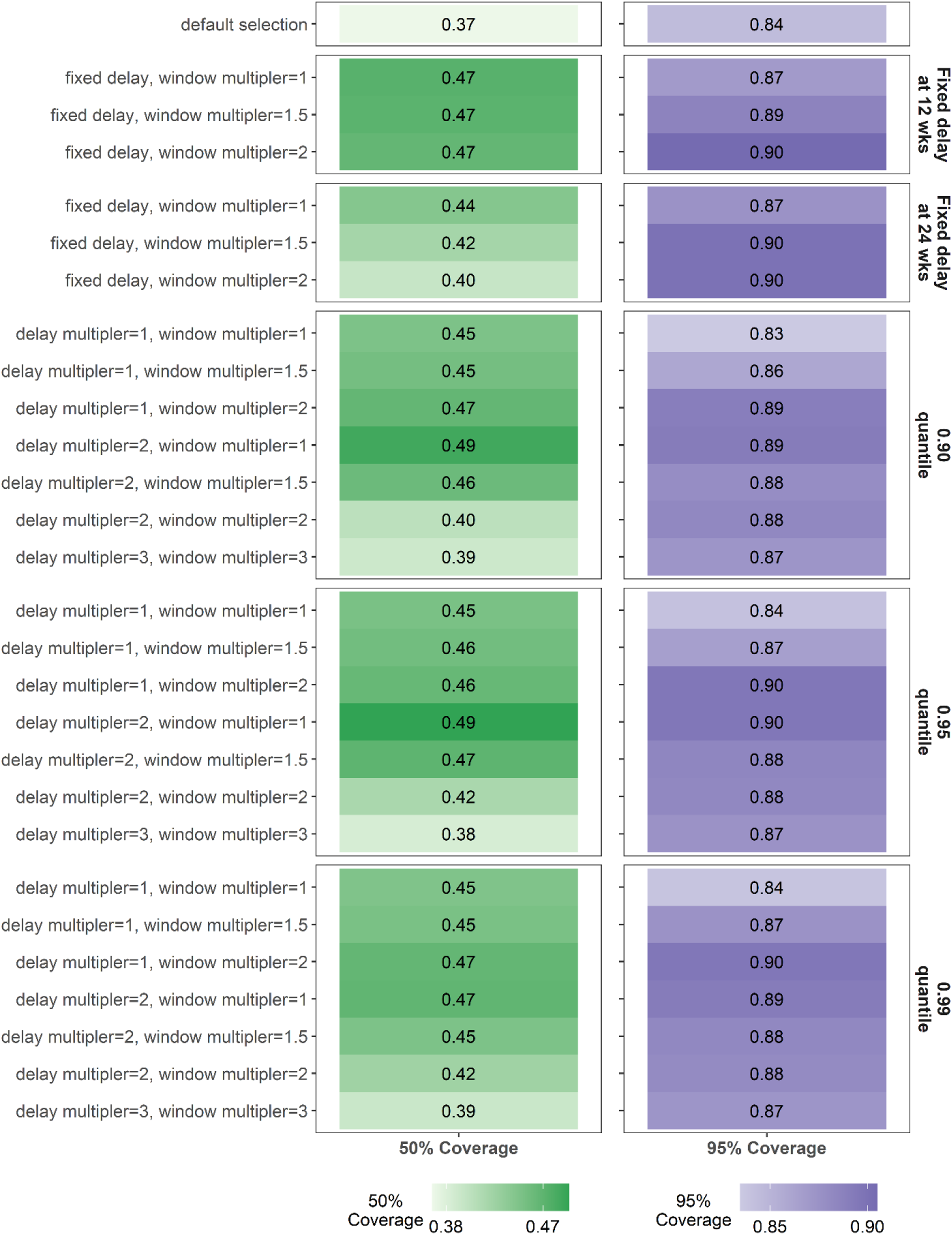
Mean 50% prediction interval coverage and 95% prediction interval coverage per parameter set. The hue of the mean prediction interval coverage is darker as coverage increases. Nowcasts classified as “Failed” were removed.

### Failed nowcasts

Even with the most robust dynamic maximum delay and training window parameters, 19% of forecasts were classified as “failed.” Of the “failed” nowcasts, 12% had observed values within the 95% PIs for two or three of the last three nowcasted dates, reflecting uncertainty across a relatively wide range of case values. On the other hand, 6% of the nowcasts with logarithmic scores above our “failure” threshold had lower coverage, reflecting overly confident nowcasts at low case numbers, which could be problematic despite having reasonable logarithmic scores.

We, therefore, assessed *a priori* indicators of the epidemic trend and the nowcasts themselves that could indicate poor performance in real-time. We analyzed five features: the mean number of reported cases in the last three weeks; the widths of the nowcast 50% prediction intervals and the 95% prediction intervals relative to mean reported cases; permutation entropy of the entire history prior to the nowcast date; and permutation entropy of the 12 weeks prior to the nowcast date. With a random forest model that included all five features, more than 99% of nowcasts were classified correctly, and the sensitivity and specificity were greater than 99%. Along with the mean number of reported cases, the relative 95% prediction interval range and permutation entropy in the last 12 weeks were the most important determinants for predicting “failure” (Figure 4A).

**Figure 4:**
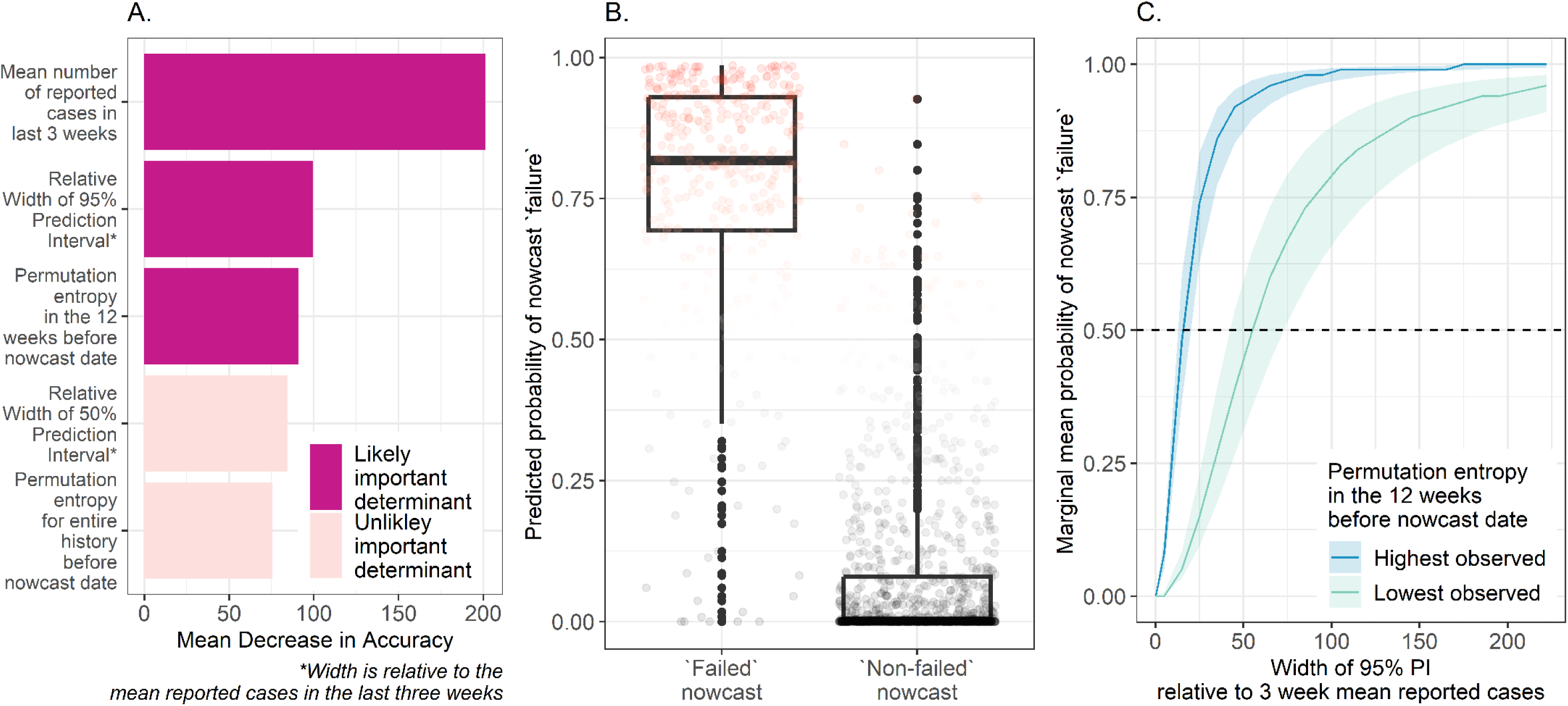
A. Variable importance, as determined by the mean decrease in accuracy, per feature from a random forest classifier. More important variables are in dark pink; B. Median and interquartile range of predicted probability of nowcast failure from the regression model for observed nowcast classification. Points in red represent predicted probabilities greater than 0.50 and points in black represent predicted probabilities less than 0.50.; C. The marginal mean probability of nowcast “failure” by width of the 95% prediction interval relative to mean cases in the last 3 weeks. Predicted values are disaggregated by the lowest and highest observed permutation entropy for the 12 weeks prior to the nowcast date. The horizontal dashed line represents the threshold values for marginal mean probability classification “failure”, where values greater than 0.50 are likely to be predicted “failures”.

To determine the indicator values predictive of nowcast failure, we fitted a logistic regression model and calculated the marginal mean probability of “failure”. The regression included three indicators: the range of the 95% prediction interval relative to recently reported cases (on the log scale), permutation entropy for the last 12 weeks, and the mean number of reported cases in the last three weeks (on the log scale). The logistic regression model fit the data well (Figure 4B), with sensitivity of 84%, specificity of 96%, and accuracy of 93% in a 20% subset of data not used to fit the original models. Because the mean case counts and log scores were strongly correlated (Pearson correlation coefficient of 0.73), we assessed the marginal mean to remove the impact of higher case numbers and focused on the associations between permutation entropy and relative width of the 95% prediction interval (Figure 4C). “Failure” was most likely with a wide 95% prediction interval and high permutation entropy. For example, when the epidemic trajectory was stable and permutation entropy was low, failure probability did not reach 50% up to a relative 95% prediction interval width of approximately 55 at the highest observed permutation entropy value (0.69). In contrast, when the epidemic trajectory was unstable and permutation entropy was high, the threshold value for the relative 95% prediction interval width was much lower, approximately 15 at the highest observed permutation entropy value (0.69).

### Model comparison

Using the optimal parameter set described above – 95% dynamic maximum delay value and a training period 1 week longer than the delay – we compared the NobBS package to two other publicly available R nowcasting packages: nowcaster, and epinowcast. Assessing nowcast performance on all dates for Idaho (relatively short delays and low variation) and Michigan (relatively long delays and high variation), average 50% prediction interval coverage was highest for NobBS (29%) and the 95% prediction interval coverage was highest for nowcaster (68%) but both were well below the nominal values (Figures 5A and B). Relative WIS was lowest for NobBS followed by nowcaster, and then epinowcast (Figure 5C). We did not apply the “failure” criteria because we could not calculate logarithmic scores for nowcasts for epinowcast.

**Figure 5:**
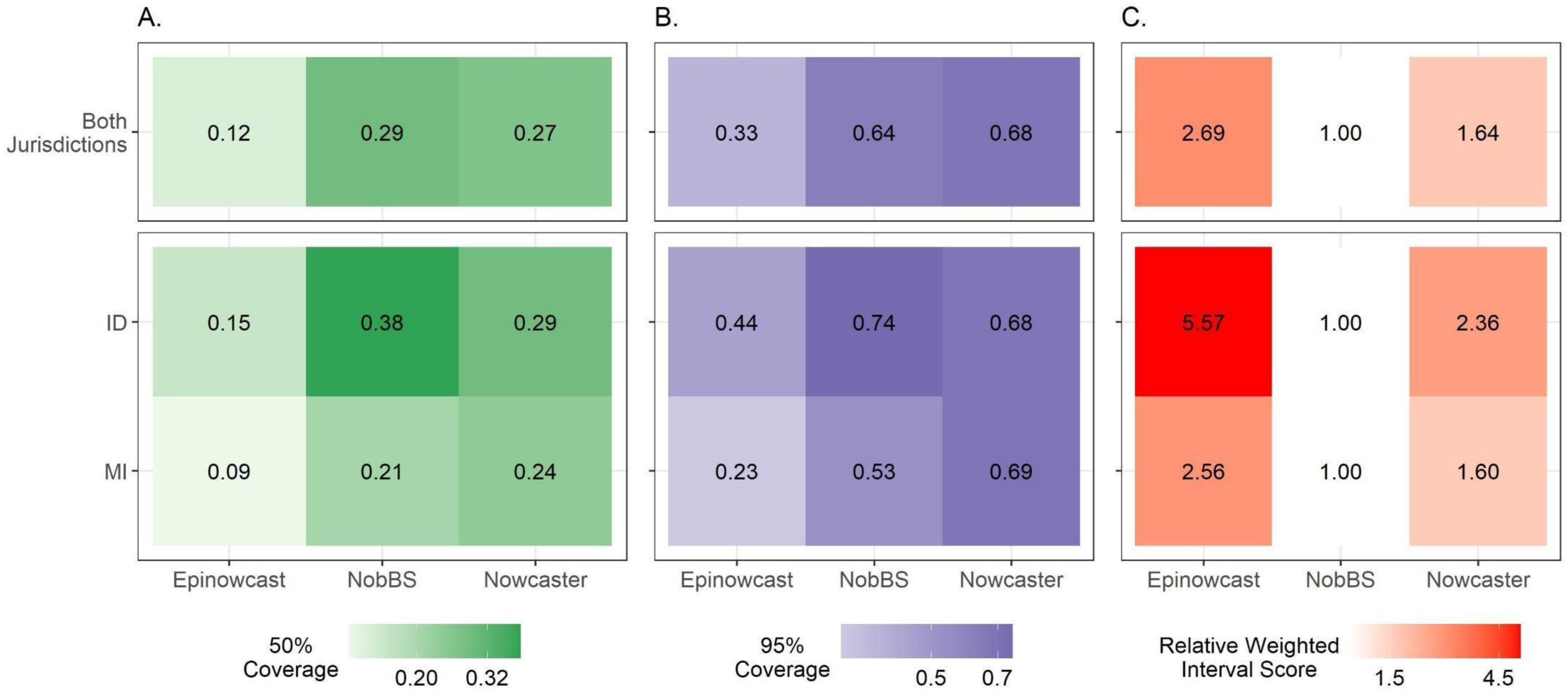
A. Mean 50% prediction interval coverage, and B. 95% prediction interval coverage per model for all runs and per jurisdiction. The hue of the mean prediction interval coverage is darker as coverage increases. C. Scaled, pairwise relative Weighted Interval Score (rWIS) (see *Methods* for description) by each model, for all runs and per jurisdiction. All models used equivalent parameters for the nowcast: a 95% dynamic maximum delay value and a training period 1 week longer than the delay. Nowcasts for all dates were included without any failure classification.

We also monitored run time and convergence for the Idaho and Michigan nowcasts. The median run time for a single nowcast was 276 seconds for epinowcast, 19 seconds for nowcaster, and 2 seconds for NobBS. We 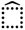found that 32% and 38% of NobBS nowcasts for Idaho and Michigan, respectively, and 5% and 13% nowcasts using epinowcast had at least one parameter or prediction with a 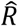 value greater than 1.01 using the default MCMC parameters (see Supplement 5). While this limited convergence, particularly with NobBS, it is a fairly stringent metric. NobBS 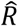 values exceeding 1.01 range from 1.02 to 1.18 and for epinowcast, 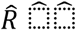 values range from 1.02 to 1.32. Nevertheless, we did not find evidence of correlation between this convergence metric (one parameter or prediction with a 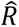 value greater than 1.01) and nowcast failure (log scores less than -9.2) for NobBS nowcasts. Overall, this adds to our confidence that the impact of non-convergence by this metric is likely minimal.

## Discussion

Our analysis provides a roadmap for nowcast implementation as a component of routine public health situational awareness by focusing on standardized, data-driven implementation choices that optimize performance. We examined performance resulting from different parameter set choices and showed that using recent data to estimate delay and training window parameters dynamically resulted in better nowcast skill than when using all historic data. For example, when the longest sets of delays were used, these models often had the poorest performance, which suggests that including too much trend data inhibited the model from capturing recent changes in trajectories. Appropriately sized delay and training windows enable the models to include important delays while also enabling adaptation as delays change. We also examined whether poor nowcasts could be predicted *a priori* and identified that the relative width of the prediction intervals and the permutation entropy of the epidemic trend can be used to guide nowcast suppression of potentially misleading nowcasts. Finally, we showed that more complex models do not significantly improve performance compared to simple models. Together, these findings help further not only the science of nowcasting but also its utility in public health decision-making.

Much of the published public health nowcasting research has focused on how to improve model performance within a specific context. While there has been some exploration similar to ours that examines the impact of sliding window size on model accuracy (22), most researchers have suggested two primary approaches for reducing error: augmenting models with additional data inputs or changing the model structure. For example, dengue and chikungunya virus nowcasts that include internet data, such as search trends on Google or Twitter, in addition to reported case counts, had lower error relative to the same models with case counts alone (23,24). Internet search data has also been shown to improve performance for influenza in ensemble nowcasts (25). Similarly, improved performance has been observed when including genomic data in COVID-19 nowcasts (26). The correlation between performance improvements and additional data, nevertheless, is not always positive for all predicted outcomes. Recent work by Klaassen and colleagues shows that augmenting COVID-19 models with wastewater surveillance data only improved nowcasts for deaths and not cases (27). Additionally, inclusion of day-of-the-week effects in models has shown some improvement in prediction interval coverage but limited meaningful impact on error metrics (8). In contrast to this body of literature, our work examines how to improve nowcasting predictions without additional data elements and across a wide range of surveillance systems.

Some researchers have suggested that nowcasting approaches should be designed for each specific public health surveillance system (11). We argue that nowcasting should be flexible in order to scale-up and be useful for many jurisdictions of different sizes. Thus, we tested nowcasting in epidemic and endemic disease surveillance systems with different reporting delay patterns to improve generalizability of our findings. We also expanded our analysis to shed light on when nowcasts may provide misleading information, which not only adds to their flexibility but also provides practical insight. Nevertheless, our approach has four main limitations. First and foremost, we examined nowcasting performance on aggregate and did not explicitly examine performance at different points in an epidemic trajectory, that is, when the trend in cases may rapidly change directions. Such an analysis may provide insight into why models with default parameters outperformed those with dynamic parameters in certain periods. Moreover, there are many ways of assessing nowcast performance, including whether the direction of the epidemic trend or specific threshold values were predicted. Nowcasting methods are generally not good at capturing changes in epidemic trajectory, and changes in the reporting delays themselves may also correlate with changes in the epidemic trend. While we did include jurisdictions with high variance in reporting delays to test the impact of parameter selection, we did not explicitly examine the relationship between changes in delay patterns and corresponding changes in epidemic trajectory. Further research is needed in this area. Second, there is no clear, objective definition of “failure”. We employed a “failure” metric that is correlated with the scale of the observed outcome; consequently, there may be some misclassification of nowcasts. Third, we observed some nowcasts that may not have converged, emphasizing the need for surveillance officers and modelers to carefully assess model performance and identify approaches to improve convergence. Finally, unlike other research, we did not test how additional data may impact model performance and aggregated daily reports to the weekly scale to smooth over weekday reporting effects. Nor did we compare predictions across different modeling approaches. Further research on predictive skill of nowcast models that combine dynamic parameter selection and augmented datasets is needed, and comparing our nowcasting approach to others used in different disciplines (such as Kalman filtering or sequential Monte Carlo) could provide additional insight.

Nowcasting implementation has been most successful when analysts closely collaborate with surveillance partners. We sought to identify options to build more robust nowcast predictions when nowcasts are implemented in real-time, and assessing performance for individual nowcasts becomes untenable. Towards this end, our findings yield three main recommendations:

1. Prior to running models, we recommend that batch reporting patterns be identified with surveillance experts. Implications of poor nowcast skill under batch reporting conditions should be discussed, and suppressing nowcasts should be considered.
2. Programmatically setting maximum delay parameters dynamically for each week based on the distribution of onset-to-report delays for the most recent reported data resulted in the best performance. A fixed maximum delay of 12 weeks also outperformed the default parameters. The training window parameter can be set to the maximum delay value plus one week.
3. We recommend that a system be implemented to suppress nowcasts likely to fail. This involves programmatic evaluation of the relative 95% prediction interval width and permutation entropy in the most recent 12 weeks. As a conservative approach, when the ratio of the width of the 95% prediction interval to the mean reported cases in the most recent three weeks is less than 10, the nowcast estimates are likely to be stable. This approach, like using a coefficient of variation, highlights a reasonable dispersion of the prediction relative to the mean reported cases.

## Data Availability

COVID-19 case data are available upon request at https://data.cdc.gov/Case-Surveillance/COVID-19-Case-Surveillance-Restricted-Access-Detai/mbd7-r32t. Dengue case data for 2010 are available from https://github.com/sarahhbellum/NobBS/blob/master/data/denguedat.RData and dengue case data from 2011-2014 were obtained from the Puerto Rico Department of Health. R code is available in a public repository (https://github.com/cdcepi/Nowcasting_scale_up_analysis).

## CDC disclaimer

The findings and conclusions in this report are those of the authors and do not necessarily represent the official position of the U.S. Centers for Disease Control and Prevention.

## Declarations

### Ethics approval and consent to participate

Not applicable

### Consent for publication

Not applicable

### Clinical trial number

not applicable

### Authors’ contributions

VKL and MAJ designed the research question and analytical approach, with input from LB and CC. VKL conducted all analyses and wrote the first version of the manuscript with contributions from MAJ. All authors reviewed, edited, and approved the final manuscript.

## Footnotes

1§ See e.g., 45 C.F.R. part 46, 21 C.F.R. part 56; 42 U.S.C. §241(d); 5 U.S.C. §552a; 44 U.S.C. §3501 et seq.

