## Supplement for "A roadmap to account for reporting delays for public health situational awareness – a case study with COVID-19 and dengue in United States jurisdictions"

**Authors:** *Velma K. Lopez[1], Leonardo S. Bastos[2], Cláudia T. Codeço[2], Michael A. Johansson[1]*
**Affiliations:**
1. Centers for Disease Control and Prevention, Dengue Branch, San Juan, Puerto Rico
2. Fundação Oswaldo Cruz, Programa de Computação Científica, Rio de Janeiro, Brasil

**Supplemental Figures and Tables**

**Supplemental Figure 1: Median and interquartile range of weekly reporting delays per jurisdiction.** Jurisdictions are grouped into four categories based on the mean and standard deviation of the reporting delays: high mean and high standard deviation, low mean and high standard deviation, low mean and low standard deviation, and other combinations (which were excluded). Jurisdictions with an asterisk were randomly selected for inclusion in the analysis.

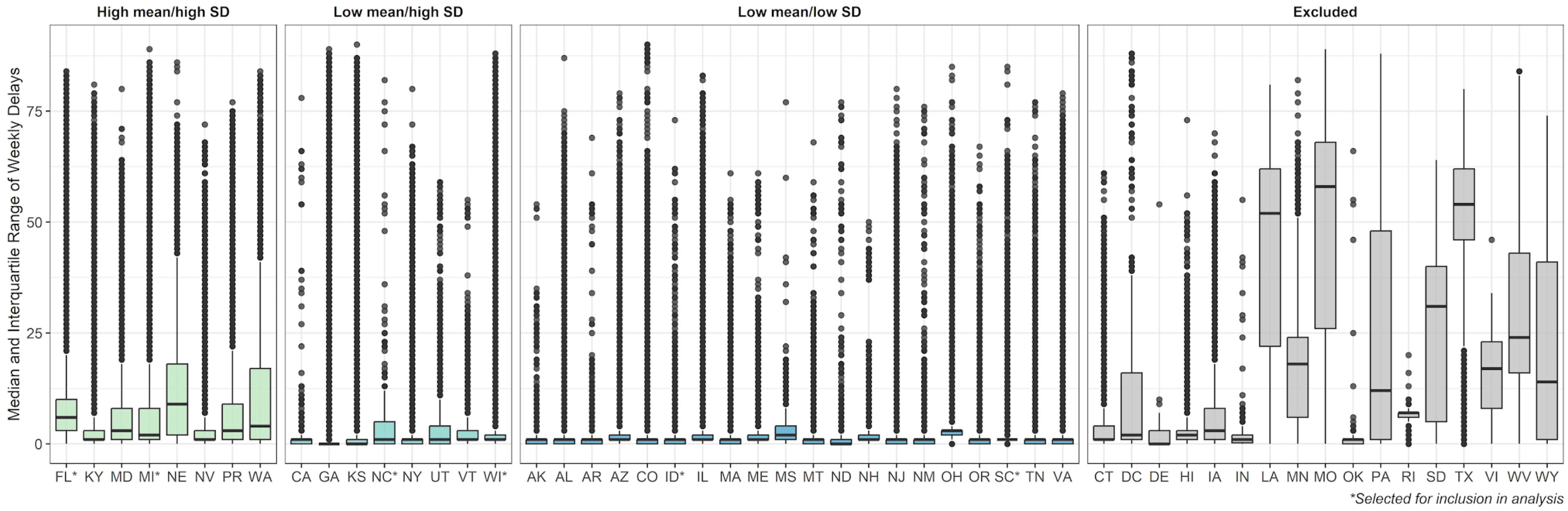

**Supplement 2**

**Supplemental Figure 2.1**: Percent of nowcasts where the model with default parameter outperformed the models with dynamic parameters, by jurisdiction and dynamic parameter set.

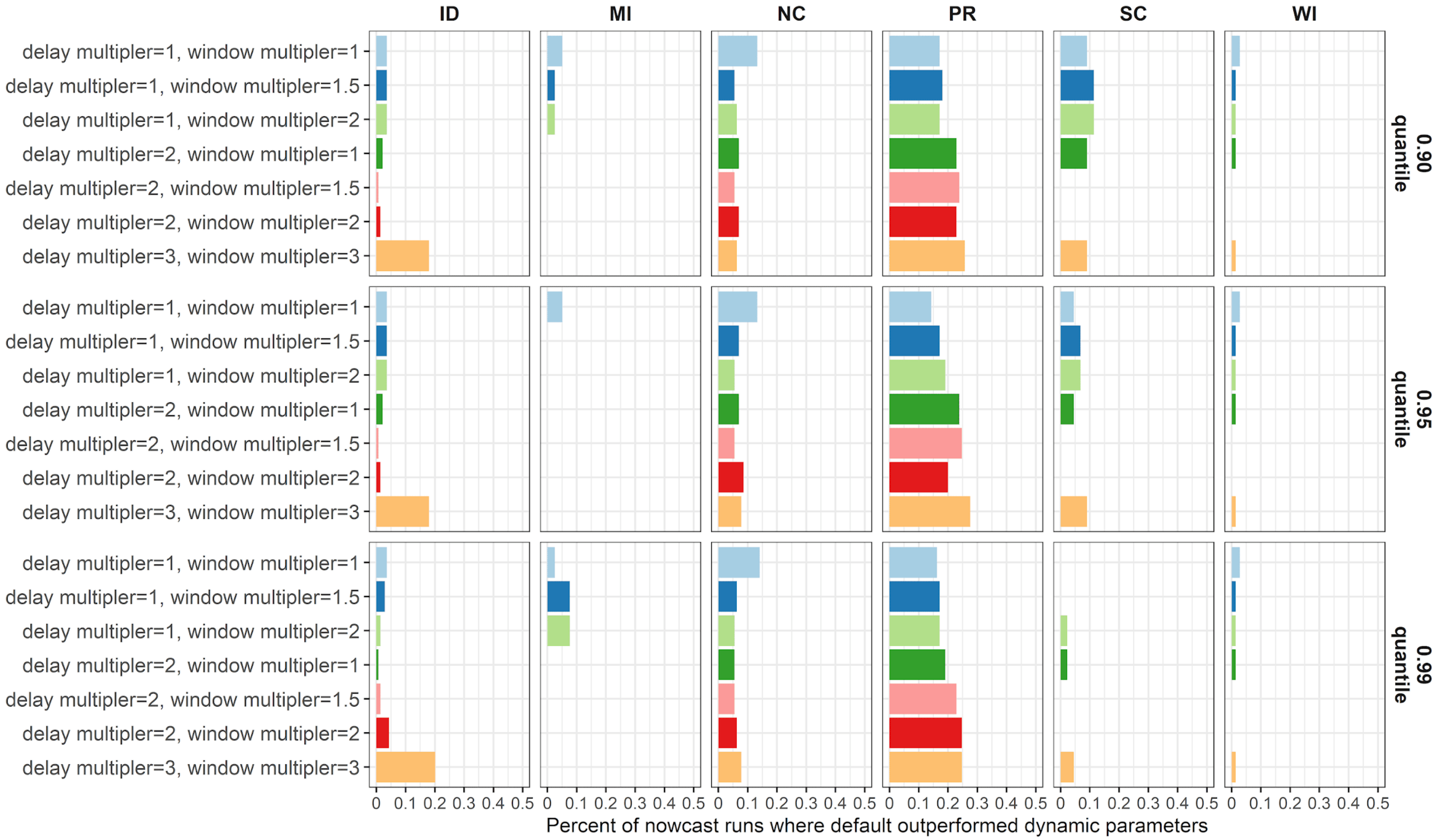

**Supplemental Figure 2.2: Unadjusted mean log scores over time per jurisdiction.** The horizontal dashed line represents the threshold for a “failed” nowcast. Log scores below this threshold were excluded from the evaluation. In each plot, the parameter set is presented in a distinct color, with the default (in black) presented as a reference for every parameter group.

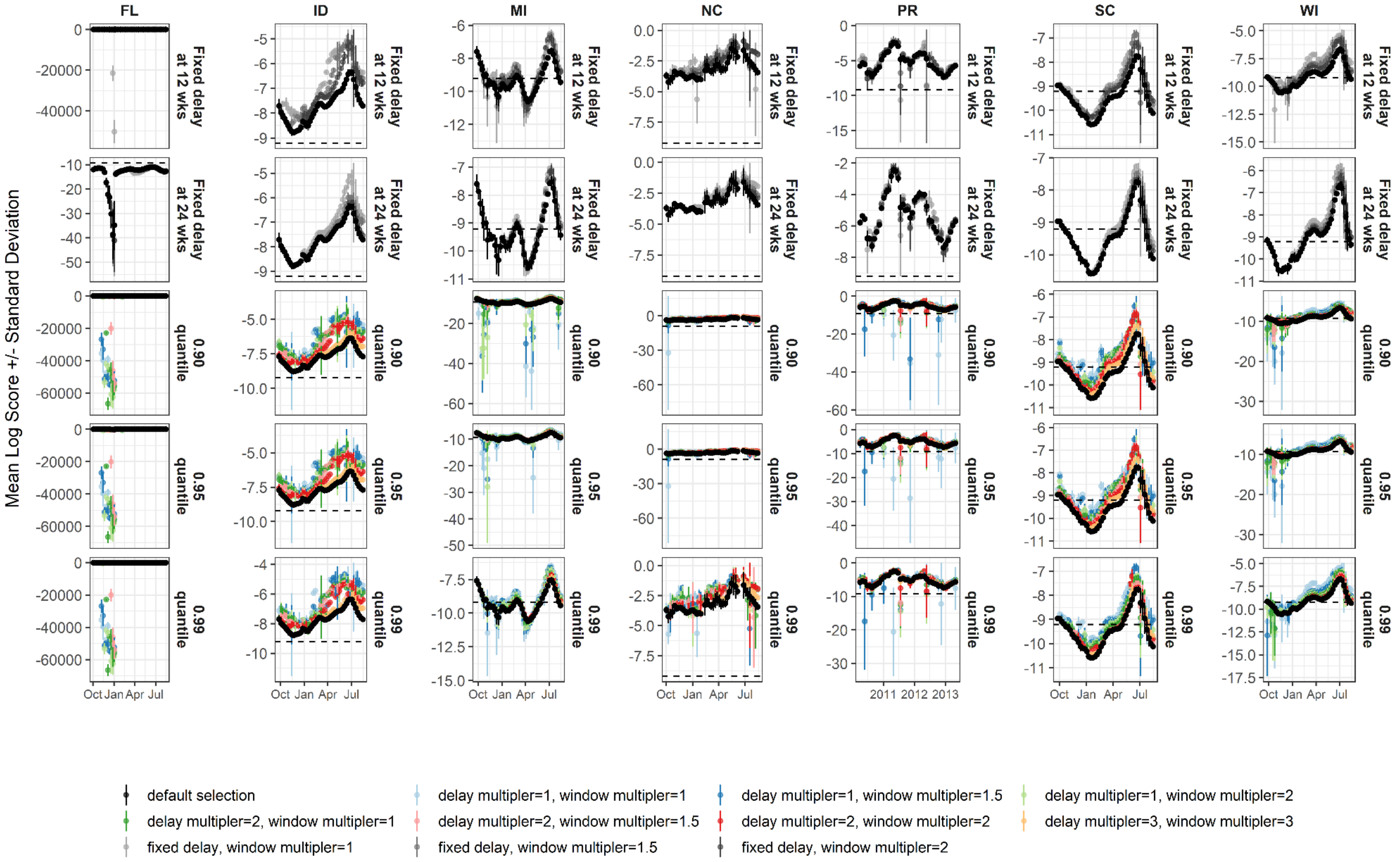

**Supplemental Figure 2.3. Jurisdictional log scores per parameter set.** A. The percent of nowcasts with failed log scores, which were excluded from the evaluation; B. Median log score (as a vertical line), interquartile range log scores (as a horizontal bar), and mean log score (as a yellow diamond); C. The parameter set mean log score relative to mean log score of the fixed delay of 12 weeks with 1 window multiplier. In each plot, the parameter set is presented in a distinct color.

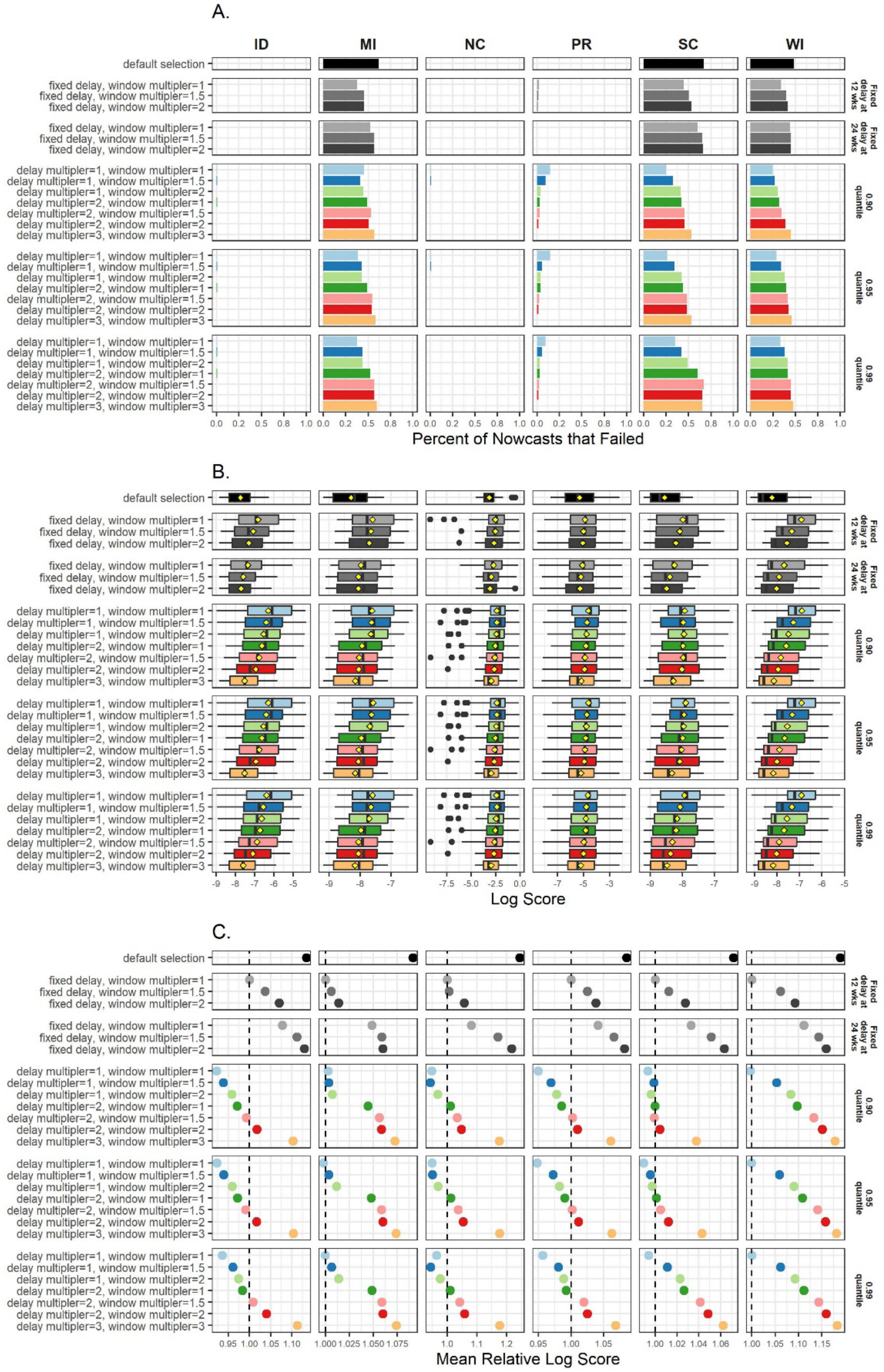

**Supplemental Figure 2.4: Jurisdictional mean 95% prediction interval coverage per parameter set.** The hue of the mean 95% prediction interval coverage is darker as coverage increases.

| Group | Parameter set | ID | MI | NC | PR | SC | WI |
|---|---|---|---|---|---|---|---|
| | default selection | 0.87 | 0.46 | 0.93 | 0.94 | 0.82 | 0.72 |
| Fixed delay at 12 wks | fixed delay, window multipler=1 | 0.85 | 0.64 | 0.95 | 0.90 | 0.84 | 0.86 |
| Fixed delay at 12 wks | fixed delay, window multipler=1.5 | 0.81 | 0.72 | 0.98 | 0.90 | 0.86 | 0.96 |
| Fixed delay at 12 wks | fixed delay, window multipler=2 | 0.81 | 0.87 | 0.98 | 0.90 | 0.86 | 1.00 |
| Fixed delay at 24 wks | fixed delay, window multipler=1 | 0.80 | 0.64 | 0.97 | 0.89 | 0.86 | 0.97 |
| Fixed delay at 24 wks | fixed delay, window multipler=1.5 | 0.88 | 0.59 | 0.96 | 0.93 | 0.89 | 0.96 |
| Fixed delay at 24 wks | fixed delay, window multipler=2 | 0.88 | 0.54 | 0.96 | 0.94 | 0.86 | 0.97 |
| 0.90 quantile | delay multipler=1, window multipler=1 | 0.75 | 0.56 | 0.92 | 0.93 | 0.86 | 0.83 |
| 0.90 quantile | delay multipler=1, window multipler=1.5 | 0.77 | 0.69 | 0.92 | 0.94 | 0.82 | 0.92 |
| 0.90 quantile | delay multipler=1, window multipler=2 | 0.81 | 0.85 | 0.91 | 0.92 | 0.91 | 0.96 |
| 0.90 quantile | delay multipler=2, window multipler=1 | 0.86 | 0.62 | 0.96 | 0.93 | 0.86 | 0.93 |
| 0.90 quantile | delay multipler=2, window multipler=1.5 | 0.86 | 0.56 | 0.96 | 0.90 | 0.82 | 0.94 |
| 0.90 quantile | delay multipler=2, window multipler=2 | 0.83 | 0.54 | 0.98 | 0.92 | 0.86 | 0.96 |
| 0.90 quantile | delay multipler=3, window multipler=3 | 0.88 | 0.49 | 0.95 | 0.94 | 0.86 | 0.82 |
| 0.95 quantile | delay multipler=1, window multipler=1 | 0.75 | 0.62 | 0.92 | 0.93 | 0.86 | 0.85 |
| 0.95 quantile | delay multipler=1, window multipler=1.5 | 0.77 | 0.72 | 0.92 | 0.94 | 0.82 | 0.94 |
| 0.95 quantile | delay multipler=1, window multipler=2 | 0.81 | 0.87 | 0.91 | 0.92 | 0.91 | 0.99 |
| 0.95 quantile | delay multipler=2, window multipler=1 | 0.87 | 0.64 | 0.96 | 0.92 | 0.84 | 0.96 |
| 0.95 quantile | delay multipler=2, window multipler=1.5 | 0.86 | 0.59 | 0.96 | 0.90 | 0.84 | 0.96 |
| 0.95 quantile | delay multipler=2, window multipler=2 | 0.83 | 0.54 | 0.98 | 0.91 | 0.86 | 0.97 |
| 0.95 quantile | delay multipler=3, window multipler=3 | 0.88 | 0.49 | 0.95 | 0.94 | 0.86 | 0.83 |
| 0.99 quantile | delay multipler=1, window multipler=1 | 0.74 | 0.64 | 0.94 | 0.91 | 0.82 | 0.86 |
| 0.99 quantile | delay multipler=1, window multipler=1.5 | 0.80 | 0.72 | 0.93 | 0.91 | 0.86 | 0.96 |
| 0.99 quantile | delay multipler=1, window multipler=2 | 0.83 | 0.87 | 0.91 | 0.91 | 0.86 | 1.00 |
| 0.99 quantile | delay multipler=2, window multipler=1 | 0.85 | 0.64 | 0.96 | 0.91 | 0.86 | 0.97 |
| 0.99 quantile | delay multipler=2, window multipler=1.5 | 0.84 | 0.59 | 0.95 | 0.90 | 0.91 | 0.96 |
| 0.99 quantile | delay multipler=2, window multipler=2 | 0.83 | 0.54 | 0.97 | 0.90 | 0.89 | 0.97 |
| 0.99 quantile | delay multipler=3, window multipler=3 | 0.87 | 0.49 | 0.95 | 0.96 | 0.84 | 0.83 |
| | | 95% Coverage | 95% Coverage | 95% Coverage | 95% Coverage | 95% Coverage | 95% Coverage |

95% Coverage: 0.5 0.7 0.9

**Supplemental Figure 3: Feature correlation.** Pearson correlation coefficient for all features included in the random forest model. Dark colors reflect stronger correlations.

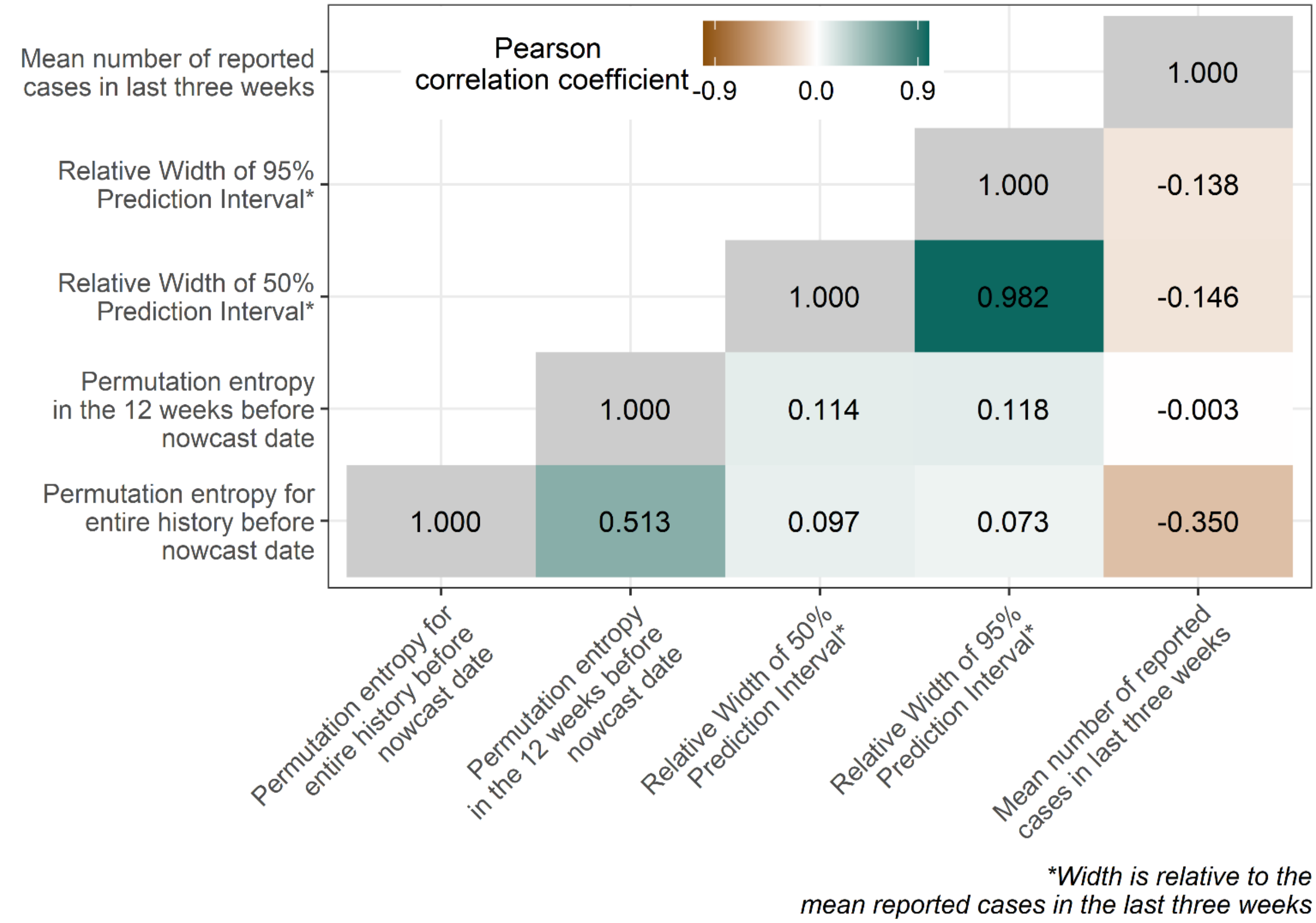

**Supplemental Table 4**: Differences and similarities in underlying model structure between NobBS, Nowcaster, and Epinowcast

| | NobBS | Nowcaster | Epinowcast |
|---|---|---|---|
| *Time scale* | Daily or weekly | Daily or weekly | Daily |
| *Key parameters* | | | |
| Maximum delay | User-defined; defaults to the entire history - 1 | User-defined; defaults to 15 weeks | User-defined; defaults to 20 days |
| Window period | User-defined; defaults to the entire history | User-defined; defaults to 30 weeks | Not changeable; uses entire history |
| *Model of reporting the distribution of delays* | | | |
| Delays are modeled as: | As a probability | As a function of time | As a probability |
| Variance around delays are given a: | Weakly informative Dirichlet prior (0.1) | Second-order random walk | Half-normal prior (0.1) |
| Distribution | lognormal | lognormal | User-defined; supports no distribution, lognormal, gamma, exponential, log-logistic, and lognormal, with lognormal default |
| *Model of observed counts* | | | |
| Underlying model structure: | Negative binomial or Poisson model | Negative binomial model | Negative binomial or Poisson model |
| Variance around time is given: | First-order random walk | First or second-order random walk | First-order random walk |
| *Model of expected counts* | * | * | Exponential growth rate model on the log scale |
| *Model of difference in reporting delay distribution by date of report* | * | * | A random effect for weekday or weekly reporting effects |
| *Inference by* | Markov chain Monte Carlo (MCMC) via JAGS | Integrated nested Laplace approximation (INLA) | MCMC via CMDStanR |

** not included in the model framework*

**Supplement 5: $\hat{R}$ > 1.01 for epinowcast and NobBS and run times for all models.** A. The proportion of all nowcast runs, per jurisdiction, where at least one prediction had an $\hat{R}$ value > 1.01; B. The distribution of $\hat{R}$ values > 1.01 per jurisdiction; C. The distribution of total run time in seconds, per model (n=94).

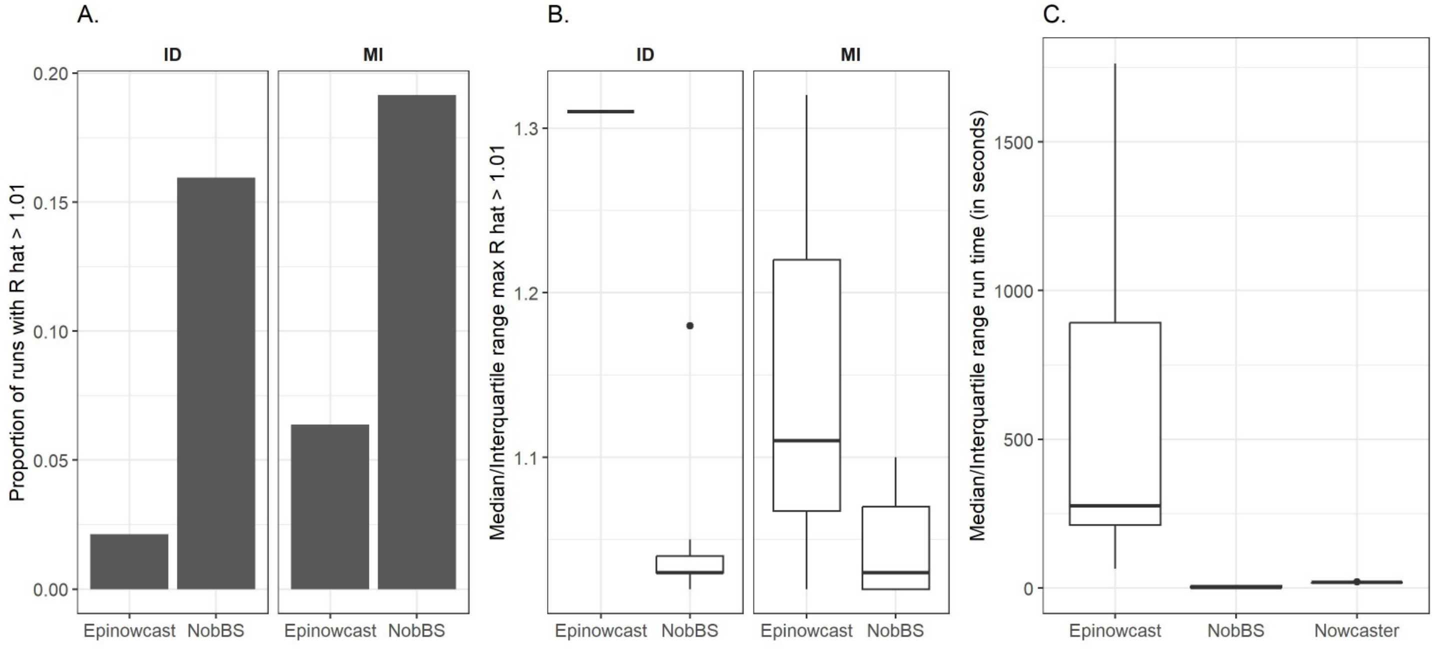
